# Arg4810Lys Mutation in *RNF213* among Eastern Indian Non-MMD Ischemic Stroke Patients: A Genotype-Phenotype Correlation

**DOI:** 10.1101/2023.05.30.23290718

**Authors:** Dipanwita Sadhukhan, Parama Mitra, Smriti Mishra, Arunima Roy, Gargi Podder, Biman Kanti Ray, Atanu Biswas, Subhra Prakash Hui, Tapas Kumar Banerjee, Arindam Biswas

## Abstract

**Introduction:** *RNF213* mutations have been reported mostly in Moyamoya Disease (MMD) with varying frequencies across different ethnicities. However, its prevalence in non-MMD adult-onset Ischemic Stroke is still not well explored.

**Aims & Objectives:** This present study thus aims to screen the most common *RNF213* variant (Arg4810Lys, among East Asians) in the Eastern Indian non-MMD Ischemic Stroke patients and correlate it with long-term progression and prognosis of the patients. The subjects were analysed for this variant using PCR-RFLP and confirmed using Sanger sequencing method.

**Result & Conclusion:** We have identified Arg4810Lys variant among eleven young-onset familial Ischemic Stroke patients in heterozygous manner. A positive correlation of the variant with positive family history (P = 0.001), earlier age-at-onset (P = 0.002), history of recurrent stroke (P = 0.015) was observed. However, the carriers showed better cognitive performances in memory (P = 0.042) and executive function (P = 0.004). Therefore, we can conclude that Arg4810Lys/*RNF213* - a pathogenic variant for young-onset familial Ischemic Stroke with higher incidence of recurrent events unlike in MMD cases, have no additional impact on cognition among Eastern Indians.

## 1. INTRODUCTION

Ischemic stroke (IS) is a leading cause of death and disability across the world. A sudden disruption in the blood flow to the brain, due to arterial stenosis or a clot in an artery carrying blood to the brain deprives the brain of oxygen and glucose resulting into multiple neuropathological events including mitochondrial dysregulations, neuroinflammation, impaired permeability of Blood-Brain-Barrier, disturbed homeostasis and eventually cellular death, loss of synaptic transmission [1,2]. Post-stroke survivors often experience long-term difficulties including motor disabilities and cognitive/memory impairment [3]. Along with non-modifiable risk factors such as positive family history, age, sex, ethnicity, modifiable environmental factors as hypertension, diabetes mellitus, alcohol consumption, smoking, diet, physical inactivity have also been identified as potential causes of stroke [4]. There are also a number of other genetic disorders, like Moyamoya Disease (MMD), that often manifest adult-onset ischemic events as a primary symptom [4].

*RNF213*, which encodes a 591 kDa protein called Mysterin, is known to be the most common causal gene harbouring MMD-associated variants among East Asians [5]. Among the plethora of variants identified in this gene, p.Arg4810Lys is the most prevalent one, which has shown reduced angiogenic capabilities *in vitro*, while, the other rare variants (His4014Asn, Cys4023Arg, Cys4017Ser, Cys3997Tyr) in *RNF213* identified in the Caucasian population have shown impaired zinc ion coordination [6] and lipid droplet targeting [7]. Its role has also been implicated in angiogenesis and inflammatory pathways [8]. Earlier reports on *RNF213* described its association with IS in the Korean population [9] and with MMD among Japanese [10], Chinese [11], Korean [12] as well as in Indian [13] populations. Thus, the given background of limited genetic studies on *RNF213* in non-MMD IS cases in international and national manner, tempted us to screen the p.Arg4810Lys variant in Eastern Indian population, and to correlate it with long-term progression and prognosis of the patients.

## 2. MATERIALS AND METHODS

### 2.1 Study Subjects

A total of 119 non-MMD East-Indian Ischemic stroke patients were recruited for this study. These cases were examined at Bangur Institute of Neurosciences (BIN), Kolkata and National Neuroscience Centre, Kolkata. In addition, 100 unrelated healthy controls (mean age, 58.3 ± 9.2 years) were selected from the same ethnic background. The demographic details of the subjects are described in Table I.

**Table 1:** Clinical characteristics of patients with and without *RNF213* (p.Arg4810Lys) mutation.

|  | **With Arg4810Lys**<br>**mutation (n = 11)** | **Without Arg4810Lys**<br>**mutation (N = 108)** | **P - value** |
| --- | --- | --- | --- |
| Age at Onset | **41.9 ± 11.15** | **55.49 ± 11.29** | **\*0.002** |
| Gender, Male | 10 (91%) | 84 (77.78%) | **0.454 |
| Hypertension Positive | 8 (73%) | 72 (66.67%) | **1.000 |
| Type-2 Diabetes Positive | 4 (36%) | 44 (41%) | **1.000 |
| Positive Family History | **11 (100%)** | **54 (50%)** | **\*\*0.001** |
| Recurrent event | **7 (64%)** | **28 (26%)** | **\*\*0.015** |
| BMSE | 25.67 ± 2.91 | 23.82 ± 6.02 | *0.815 |
| GDS | 11.44 ± 6.52 | 11.74 ± 6.03 | *0.846 |
| Attention | 12.37 ± 3.20 | 9.36 ± 5.09 | *0.117 |
| Orientation | 9.13 ± 0.99 | 8.20 ± 2.29 | *0.422 |
| Memory | **42.44 ± 8.00** | **33.28 ± 12.72** | **\*0.042** |
| Language | 32.62 ± 5.97 | 31.10 ± 10.52 | *0.835 |
| Visuospatial skills | 6.55 ± 2.00 | 5.20 ± 3.32 | *0.192 |
| Calculation | 3.67 ± 2.29 | 3.75 ± 2.17 | *0.991 |
| Executive function | **13.11 ± 3.40** | **7.88 ± 5.01** | **\*0.004** |

BMSE: Bengali version of MMSE; GDS: Geriatric Depression Scale
\* Mann Whitney *U* test (P-value); \*\*Fisher exact test (P-value)

### 2.2 Screening of Arg4810Lys Mutations

Peripheral blood samples were collected from study subjects and Genomic DNA was isolated. The targeted region harboring the nucleotide change was amplified by PCR, digested with Hpy188I (New England Biolabs, USA), conditions specified by the manufacturer and electrophoresed on a 7% polyacrylamide gel. The nucleotide change identified by RFLP analysis was also confirmed by bi-directional Sanger Sequencing methods.

### 2.3 Statistical Analysis

Demographic, clinical features and cognitive parameters were analyzed using Mann Whitney *U* test and Fisher exact test. The probability level of ≤ 0.05 was considered statistically significant.

## 3. RESULTS

### 3.1 Genetic study

The p.Arg4810Lys variant of *RNF213* which is associated with the Moyamoya disease & Ischemic Stroke in the Asian population, was screened in our study cohort consisting of 119 non-MMD ischemic stroke patients and 100 ethnically matched control. Among them, eleven patients (°9.24%) were found to be heterozygous for this variant. However, none of the control individuals appeared to be carrier for this variant.

### 3.2 Comparison of risk factors

A number of risk factors for ischemic stroke such as, positive family history, hypertension, diabetes mellitus, total cholesterol, triglyceride levels, etc. were compared between the carriers and non-carriers of p.Arg4810Lys variant in our study cohort. Our data suggests that, p.Arg4810Lys segregates in familial manner (P = 0.001) and influences early age at onset (41.9 years vs 55.49 years; P = 0.002). However, the hypertension, diabetes mellitus status and gender bias did not show any significant difference between these two groups.

### 3.3 Genotype-phenotype correlation

On documenting the phenotypes among these eleven mutant individuals, we did not observe phenotypic heterogeneity, except infarct location. Out of these eleven cases, seven had Middle Cerebral Artery (MCA) territory infarcts (°63%), three Posterior Cerebral Artery (PCA) infarct (°27%) and one Internal Carotid Artery (ICA) Occlusion (°10%).

However, a higher recurrent incidence was observed in the carriers than the other group in a statistically significant manner (P = 0.015).

Next, we compared the post-stroke cognitive complications and depression, between carriers and non-carriers. Interestingly enough, the carriers of this particular variant exhibited higher memory (42.44±8.00 vs. 33.28±12.72; P = 0.042) and better executive functional abilities (13.11± 3.40 vs. 7.88± 5.01; P = 0.004) than non-carriers. Other cognitive domains like attention, orientation, language, visuo-spatial skills, calculation, showed no such differences between the two groups.

## 4. Discussion and Conclusion

In our present study, we report *RNF213* c.14429G>A (p.Arg4810Lys) variant among eleven young onset non-MMD Ischemic stroke cases with strong positive family history and higher recurrent incidences, in heterozygous manner from East Indian population. The structural, *in-vitro* and animal studies on the p.Arg4810Lys mutation, which lies in the E3 core at the C-terminal region of this E3 ligase demonstrate the inhibition of angiogenesis by inhibiting ATP hydrolysis [15]. Our genotype data are similar with a Japanese report showing a higher prevalence of this variant amongst the early-onset IS cases with anterior circulation stenosis [16], while, a study from Korea had shown association of non-MMD IS with rs37144113; c.4950G>A variant of *RNF213*, but not with rs112735431; Arg4810Lys [9]. An extended subgroup analysis in an earlier report from South India also mentioned over representation of *RNF213* c.14429G>A variant in MMD cases those were mostly from West Bengal and Tamil Nadu [13]. Although, our Indian population is ethnically and genetically diverse across different geographical locations, this variant was found to be causative for both MMD and non-MMD IS. This is in accordance with a study highlighting that variant diversity of *RNF213* may predispose distinct populations to dissimilar cerebrovascular diseases [15].

It is evident from Table 2, that presence of positive family history is a common feature for both Indian and Japanese non-MMD IS [16]. However, the limited data availability for Japanese study restricted us from further comparative analysis between two populations. Next, when we compared demographic and other post-stroke complications among the Asian reports, only the positive family history appeared to be significantly associated with both MMD and Non-MMD IS across the studied cohorts. Interestingly, the earlier Indian report on MMD is inconsistent with our current data stating positive correlation between recurrent incidents and carrier status among Indians. On the other hand, while seizure or stroke related epilepsy was less common among *RNF213* c.14429G>A carrier than non-carriers for MMD, no such trend was observed for non MMD-IS.

**Table 2:**
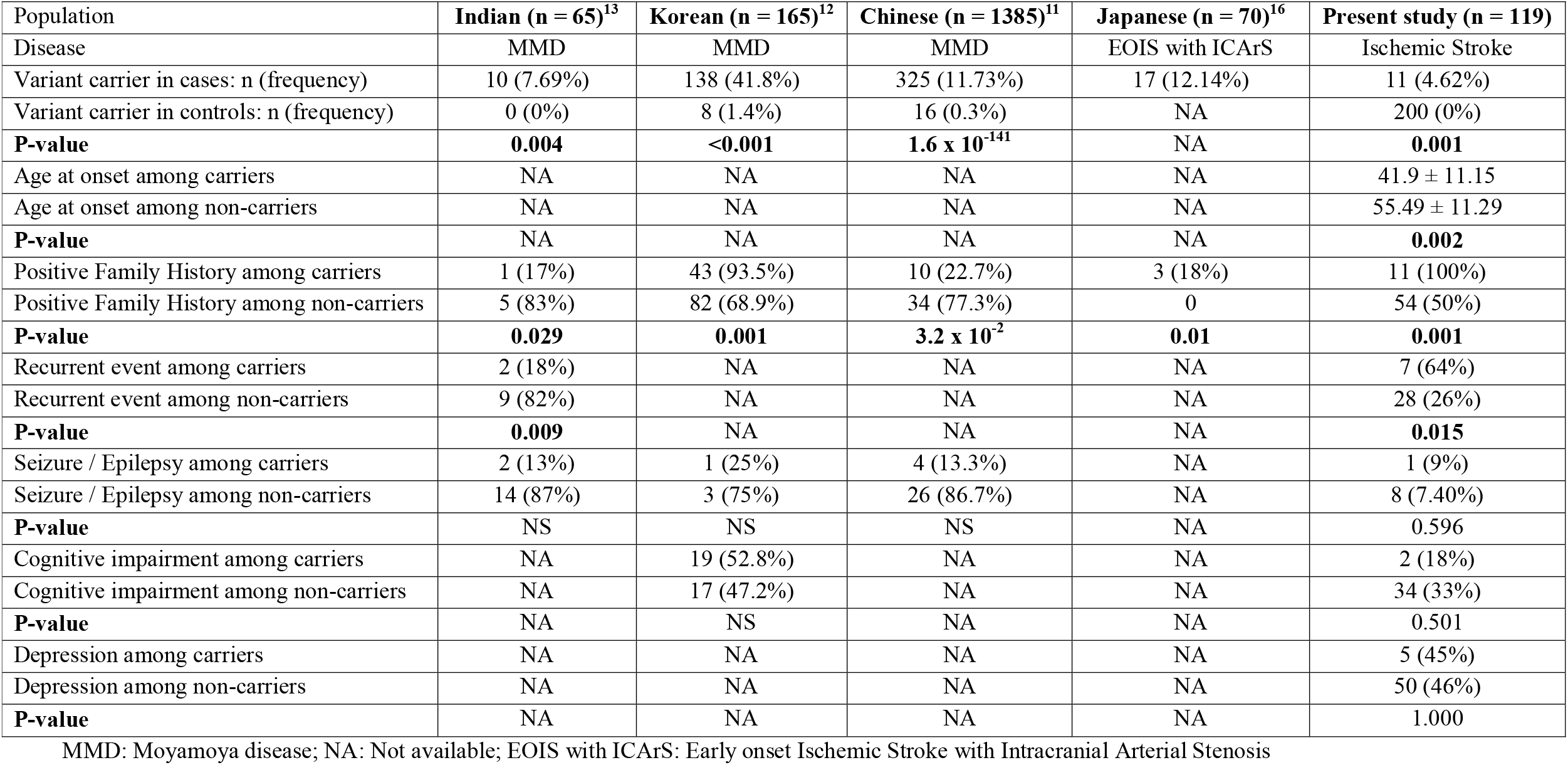
Comparison of Arg4810Lys mutation associated clinical parameters among Asians.

In general, the infarcts in the left MCA or PCA might often lead to cognitive impairment. In our study, the major locations of infract were in the MCA territory, followed by the PCA. However, we have found a better cognitive performance with respect to memory and executive functions for carriers than non-carrier while no association was found in Korean cohort of MMD cases [12]. Thus, it would be interesting to study cognitive performances in disease specific manner and investigate molecular mechanisms of RNF213 mediated post-stroke cognitive impairment in future studies. Here, although we screened the c.14429G>A variant of *RNF213* in non-MMD Ischemic stroke patients, additional variants of *RNF213* needs to be screened in a hypothesis free manner. Since there is a genetic divergence even between the two groups of same ethno-linguistic cluster among Indians, recruitment of additional samples would be warranted to strengthen our preliminary study in future. Unfortunately, here, we could not comment on the maternal or the paternal origin of the mutation in the eleven cases due to death or lack of family members.

In conclusion, to the best of our knowledge, this was the first mutation screening study from India considering *RNF213* p.Arg4810Lys variant in non-MMD Ischemic Stroke patients, along with correlation with cognitive parameters. Therefore, considering our data, genotyping for this variant might serve as a molecular diagnostic tool among familial young-onset ischemic cases with higher incidences of recurrent events.

## Data Availability

All data produced in the present study are available upon reasonable request to the authors

## Ethics statement

All procedures performed in studies involving human participants were in accordance with the ethical standards of the institutional and national research committee as well as the 1964 Helsinki Declaration and its later amendments.

## Informed consent

Informed consent from all the participants were received prior to clinical data and sample collection.

## Acknowledgement

The authors thank the patient, family members, and healthy individuals who participated in the study.

## Data Availability Statement

The data described in this study are available from the corresponding author upon reasonable request.

## Author’s contribution

Dipanwita Sadhukhan and Arindam Biswas were responsible for the concept, study design and manuscript preparation. Atanu Biswas, Biman Kanti Ray, Tapas Kumar Banerjee and Gargi Podder were responsible for the diagnosis, recruitment, clinical evaluation and blood collection for all study subjects. Parama Mitra, Smriti Mishra, Arunima Roy were responsible for experimental work and manuscript preparation. Subhra Prakashu Hui was responsible for manuscript preparation. All authors read the draft, provided their inputs and agreed on the final version of the manuscript.

